# Probing intratumoral metabolic compartmentalisation in fumarate hydratase-deficient renal cancer using clinical hyperpolarised ^13^C-MRI and mass spectrometry imaging

**DOI:** 10.1101/2024.05.06.24306817

**Authors:** Ines Horvat-Menih, Ruth Casey, James Denholm, Gregory Hamm, Heather Hulme, John Gallon, Alixander S Khan, Joshua Kaggie, Andrew B Gill, Andrew N Priest, Joao A G Duarte, Cissy Yong, Cara Brodie, James Whitworth, Simon T Barry, Richard J A Goodwin, Shubha Anand, Marc Dodd, Katherine Honan, Sarah J Welsh, Anne Y Warren, Tevita Aho, Grant D Stewart, Thomas J Mitchell, Mary A McLean, Ferdia A Gallagher

## Abstract

**Background:** Fumarate hydratase-deficient renal cell carcinoma (FHd-RCC) is a rare and aggressive renal cancer subtype characterised by increased fumarate accumulation and upregulated lactate production. Renal tumours demonstrate significant intratumoral metabolic heterogeneity, which may contribute to treatment failure. Emerging non-invasive metabolic imaging techniques have clinical potential to more accurately phenotype tumour metabolism and its heterogeneity.

**Methods:** Here we have used hyperpolarised ^13^C-pyruvate MRI (HP ^13^C-MRI) to assess ^13^C-lactate generation in a patient with an organ-confined FHd-RCC. Post-operative tissue samples were co-registered with imaging and underwent sequencing, IHC staining, and mass spectrometry imaging (MSI).

**Results:** HP ^13^C-MRI revealed two metabolically distinct tumour regions. The ^13^C-lactate-rich region showed a high lactate/pyruvate ratio and slightly lower fumarate on MSI compared to the other tumour region, as well as increased CD8+ T cell infiltration, and genetic dedifferentiation. Compared to the normal kidney, vascularity in tumour was decreased, while immune cell fraction was markedly higher.

**Conclusions:** This study shows the potential of metabolic HP ^13^C-MRI to characterise FHd-RCC and how targeting of biopsies to regions of metabolic dysregulation could be used to obtain the tumour samples of greatest clinical significance, which in turn can inform on early and successful response to treatment.

## Background

Fumarate hydratase-deficient renal cell carcinoma (FHd-RCC) is a rare but highly aggressive subtype of kidney cancer, often presenting with the clinical phenotype of hereditary leiomyomatosis and renal cell carcinoma syndrome (HLRCC)(1). The fumarate hydratase gene (*FH*) encodes the citric acid cycle enzyme which catalyses the conversion of fumarate to malate. Loss-of-function mutations in FH lead to the accumulation of fumarate, which competitively inhibits 2-oxoglutarate–dependent enzymes involved in epigenetic and gene expression regulation, driving tumorigenesis(2). Although the molecular mechanism for FHd-RCC generation is well characterised, specific clinical management guidelines remain lacking due to its rarity(3–5). Increased clinical use of cross-sectional imaging techniques has led to increased detection of kidney masses, but these methods are unable to accurately differentiate benign from malignant lesions or individual renal tumour subtypes, and provide only limited information on tumour aggressiveness including vascular enhancement and stage(6). Consequently, conventional imaging is insufficient to definitively guide patients on the nature of the tumour and their optimal treatment(7). Tumour biopsy is currently the gold-standard method of confirming the diagnosis prior to the treatment decision, but has limitations including test accuracy, invasiveness, complications, the potential for undergrading heterogeneous lesions due to sampling error, and the significant length of time required to obtain the results(8,9). New imaging approaches are urgently needed to facilitate early-stage tumour detection and screening for individuals at risk.

FH loss impairs cellular oxidative respiration prompting metabolic reprogramming to meet cellular energy demands. This reprogramming includes the upregulation of aerobic glycolysis, also known as the Warburg effect(10,11). Moreover, fumarate accumulation stabilises hypoxia-inducible factor 1α (HIF1α), an oncogene that activates transcriptional programs for angiogenesis and glycolysis(12). While [^18^F]fluorodeoxyglucose (^18^F-FDG) positron emission tomography (PET) is a well-established clinical technique for detecting these metabolic alterations, the technique is not recommended as a routine imaging tool in RCC due to renal excretion of the tracer as well as the potential risks of ionising radiation(13). Alternatively, proton MR spectroscopy (^1^H-MRS) can be used to non-invasively detect the accumulation of fumarate in FHd-tumours *in vivo* due to its high endogenous concentration, but it lacks the sensitivity to detect other metabolites present at lower concentrations(14–17). Hyperpolarised ^13^C-MRI (HP ^13^C-MRI) is an emerging clinical imaging technique that enables the non-invasive quantification of metabolism in tissues, particularly pyruvate-to-lactate conversion, which is suggestive of the Warburg effect(18,19). In human kidney cancer, HP ^13^C-MRI has been used to detect elevated lactate labelling in higher-grade clear cell RCC (ccRCC) tumours(20,21), and holds promise for revealing metabolic intratumoral heterogeneity(22).

Given the metabolic nature of the driver mutation in FHd-RCC, this subtype is of particular interest for the exploration of metabolic heterogeneity. Here we present the first application of HP ^13^C-MRI to detect the secondary effects on aerobic metabolism and intratumoral metabolic heterogeneity in FHd-RCC. We compare HP ^13^C-MRI to conventional ^1^H-MRI including measurements of cell density and vascularity, and the detection of the oncometabolite fumarate using ^1^H-MRS. We demonstrate how post-nephrectomy tissue analyses can be used to validate the imaging findings and provide insight into metabolic phenotypes within the tumour. Furthermore, using spatial metabolomics technologies(23–25) we examine how metabolic compartmentalisation correlates with changes in the tumour microenvironment (TME).

## Methods

### Ethics

This study was performed in accordance with Declaration of Helsinki. The participant gave written consent for ethically approved studies MISSION Substudy in Renal Cancer (REC: 15/EE/0378), ARTIST (REC: 20/EE/0200) and EMKC (REC: 19/EE/0161).

### HP ^13^C-MRI

The ^13^C-pyruvate injection was prepared as described previously(20,26). HP ^13^C images were acquired on a 3 T MR system (MR750, GE Healthcare, Wisconsin, USA) using a clamshell ^13^C transmit coil and ^13^C-tuned 8-channel array receive coil (Rapid Biomedical, Rimpar, Germany). At 12 s after the start of injection, an IDEAL spiral CSI sequence was used to acquire 5 slices, each 3 cm thick with 3 mm gaps and with the following parameters: 340×340mm^2^ FOV with matrix size 40×40, temporal resolution 4 s, 20 timepoints (repetition time 0.5 s), reconstructed in-plane resolution 128×128 and true voxel resolution 17×17×30mm^2^.

Reconstruction of HP ^13^C-MRI data was performed using the in-house MATLAB (MathWorks) script. Metabolite images from individual timepoints were summed together, and the complex imaging data from the 8-channels of the abdominal coil were combined using a singular value decomposition approach. The apparent pyruvate-to-lactate exchange rate constant, *k*_PL_, was calculated based on the frequency-domain modelling of Khegai *et al.* (2014)(27). Masks containing only voxels where the SNR in a summed image of the AUC of lactate + pyruvate was ≥5 were created to avoid voxels with a poor fit. DICOM format metabolite images were written for each slice, including summed ^13^C-pyruvate, summed ^13^C-lactate, and normalised ^13^C-lactate/pyruvate ratio (LAC/PYR), where normalisation denotes division of ^13^C-lactate by the summation of ^13^C-lactate + ^13^C-pyruvate.

Analysis of metabolite maps was performed in OsiriX Lite v.12.0.3 (Pixmeo SARL, Switzerland). Regions of interest (ROIs) were drawn manually around tumour and normal-adjacent kidney parenchyma on manually co-registered axial T_1_w LavaFlex images and propagated to metabolic maps. Mean and standard deviation of noise was measured in an extracorporeal region of the image. SNR of a metabolite (SNR_metabolite_) was calculated within each ROI based on the following Equation 1:

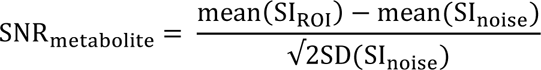

Equation 1: SNR_metabolite_ equation. SI_ROI_ = signal intensity in the ROI, SI_noise_ = signal intensity in the noise ROI.

### Physiological ^1^H-MRI

Following the HP ^13^C-MRI, the physiological ^1^H-MRI was acquired immediately after by replacing the carbon coils with the 32-channel cardiac array ^1^H coil (GE Healthcare, Wisconsin, USA) and imaging the study participant in the same 3 T MRI scanner. Protocol included T_1_w, T_2_w, IVIM-DWI and DCE-MRI sequences as described previously(20). Images were processed using proprietary (GE Healthcare) and in-house developed MATLAB-implemented software to extract the biomarker maps. Specifically, IVIM-DWI data was motion corrected across all b-values, and maps of the diffusion coefficient (*D*_0_) were computed based on non-linear fits to multi-value diffusion images.

DCE-MRI data was processed in the MiStar program (Apollo Medical Imaging Technology, Melbourne, Australia) to calculate dynamic maps of gadolinium concentration based on extended Tofts model(28) with a population-derived arterial input function (AIF). For the latter, the Fritz-Hansen AIF(29) was used, with values after the first pass extrapolated from the Weinmann function(30), scaled accordingly.

For quantification, the acquired maps underwent resampling and manual registration to the axial T_1_w LavaFlex images using ITK-SNAP 3.8 software (University of Pennsylvania, USA). ROIs of tumour and normal-adjacent kidney parenchyma were drawn in OsiriX Lite v.12.0.3 (Pixmeo SARL, Switzerland) and propagated to registered maps. Quantitative parameters were calculated from voxel intensities and reported as median (interquartile range).

### ^1^H-MRS

During the same imaging session as the conventional ^1^H-MRI, a single PRESS-localized rectangular voxel (1.7 × 1.5 cm^2^ in plane, 2.0 cm thick) was prescribed within the inside corners of the tumour. Acquisition parameters were: TE 144 ms, TR 2000 ms, number of averages 8, spectral width 5000, 4096 points, 8-step phase cycle, 64 scans with water suppression and 16 without. The choline peak at a chemical shift of 3.22 ppm, and the full width at half maximum height of the water peak in Hz, served as data quality metrics. The fumarate peak was expected at a chemical shift of 6.54 ppm. The SAGE (GE Healthcare) spectroscopy analysis software was used to reconstruct, analyse, and display spectra.

### Post-nephrectomy tissue collection

Nephrectomised kidney was halved along its superior-inferior axis, preserving the renal pelvis for diagnostic evaluation. Tissue sampling was performed from viable tumour regions based on differences in the HP ^13^C-MRI signal intensity, as well as from adjacent normal-appearing kidney parenchyma. Tissue samples were cut in half where one part was immediately flash frozen (FF) and the other part formalin-fixed and paraffin embedded (FFPE). Six samples were collected and analysed from the medial and lateral tumour portions as assessed by an experienced uropathologist (A.Y.W.).

### Targeted Next Generation Sequencing

DNA was extracted from fresh frozen specimens using the Qiagen AllPrep DNA extraction kit and following the manufacturer’s protocol. Libraries were prepared using a custom gene panel (total size: 1.455 megabases) from Twist Biosciences using the manufacturer’s protocol and were sequenced on Illumina NextSeq2000. All exons of 350 genes and flanking sequences (+/− 20 bp) are targeted with this panel. Gene list can be found in Suppl. Table 6.

Sequencing data was analysed using an in-house bioinformatics pipeline, which utilises the following main algorithms: SNVs/INDELS following GATK4/MuTect2 best practices from the Broad Institute. Non-CNV structural variations with EMBL’s Delly2 following best practices. The minimum coverage to call variants was 100X, and the targeted coverage was 500x. The limit of detection of the assay was 3% variant allele frequency. For variant filtering the following criteria was applied: allele frequencies in population databases e.g. (GnomAD, ExAC): Variants with AF> 0.002 which are reported as benign or likely benign in ClinVar can be excluded. Variants with AF> 0.01 can be excluded without previous benign classification.

An algorithmic amalgamation of 30 computational normalised scoring systems (Polyphen2_HDIV, Polyphen2_HVAR, VEST3, fathmm.MKL_coding, SIFT, MutationAssessor, GERP++, phyloP100way_vertebrate, phyloP20way_mammalian, LRT, MutationTaster, FATHMM, PROVEAN, CADD, DANN, MetaSVM, MetaLR, integrated_confidence_value, integrated_fitCons, phastCons100way_vertebrate, phastCons20way_mammalian, SiPhy_29way_logOdds, Eigen.raw, M.CAP, REVEL, MutPred, GenoCanyon, Truncation, Homozyg and XAcorrect). Any value greater than 0.60 was considered as of pathogenetic relevance.

### Copy Number Variation (CNV) analysis

CNVs were called using GATK4 following Broad Institute’s best practices. Log2 copy ratio values, which indicate changes in copy number relative to a neutral sample, were calculated with GATK4 using normalised read coverage values across the tumour sample. The mean log2 copy ratio value for each modelled segment was scaled using the pathology estimated tumour content to allow for estimation of absolute copy number. Although CNV analysis was performed, the algorithm may not detect complex deletions, duplications, or large genomic rearrangements.

### Tumour Mutational Burden (TMB)

Tumour mutational burden (muts/Mb) was calculated by dividing the number of eligible variants by the targeted sequencing region. Eligible variants are synonymous and non-synonymous somatic coding region mutations (SNV and indels) with a depth of ≥ 200 and a VAF of ≥ 5%. Since the target regions of the panel were designed to capture genes known to be frequently mutated in cancer, known somatic variants within the COSMIC database are also removed so as not to inflate TMB estimates.

### RNAseq and analysis

RNA was extracted from fresh frozen samples using the Qiagen AllPrep RNA extraction kit following the manufacturer’s protocol, and the sequencing was performed on Illumina NextSeq 500, using the high-output 75 sequencing kit. Quality control and mapping were performed using the FastQC(31) and STAR aligner(32). The RNA count data was generated through FeatureCounts(33) based on GRCh38,and normalised using the R package DESeq2 (version 1.40.1)(34). BayesPrism(35) was applied to estimate cellular composition of the tumour based on the adult kidney reference derived from normal kidneys of patients with RCC(36). Collation of major lineages (such as epithelial, endothelial, fibroblast, lymphoid and myeloid portions) was applied to minimise batch affects and inability to discern subtly different cell subtypes.

### Histology and IHC

All collected FFPE biopsies were cut at 3 μm on glass slides for staining with H&E and were assessed by a uropathologist with >15 years of experience. IHC staining for MCT1, MCT4, CD31, MIB-1 and CD8 was performed on Leica’s Polymer Refine Detection System (DS9800) in combination with Bond automated system (Leica Biosystems Ltd., Newcastle upon Tyne, UK), based on a protocol described previously(20), using the antibodies detailed in Suppl. Table 7.

HALO AI v3.6.4134 (Indica Labs, Albuquerque, NM, USA) and DenseNet v2 were used to create a tissue classifier to enable automated analysis of epithelial-stromal regions separately for all IHC staining. Results were expressed as percentage of positive tissue for specific stain or as number of positive cells and the positive optical density used were: MCT1 = 0.1106; MCT4 = 0.1106; CD31 = 0.5597,0.8142,1.0027; MIB-1 = 0.2319; CD8 = 0.2655.

### Desorption Electrospray Ionisation (DESI) Mass Spectrometry Imaging (MSI)

The FF biopsies were embedded in a hydroxypropyl methylcellulose (HPMC)/polyvinylpyrrolidone (PVP) hydrogel as previously described(37). Tissue sections were taken at 10 µm thickness using a cryostat microtome (CM3050 S, Leica Biosystems, Nussloch, Germany). Sections were thaw mounted onto Superfrost microscope slides (Thermo Scientific Waltham, Massachusetts, USA), dried under a stream of nitrogen, and stored at −80°C in sealed vacuum pouches to maintain metabolite integrity. PVP (MW 360 kDa) and HPMC (viscosity 40–60 cP, 2% in H2O (20 C) were purchased from Merck (Darmstadt, Germany). Methanol, water, isopentane and isopropyl alcohol were obtained from Fisher Scientific (Waltham, Massachusetts, USA).

DESI-MSI analysis was performed on a Q-Exactive mass spectrometer (Thermo Scientific, Bremen, Germany) equipped with an automated two-dimensional-DESI ion source (Prosolia, Indianapolis, Indy, USA). The mass spectrometer was operated in negative ion mode, with a mass range m/z of 80-900, with a nominal mass resolution of 70,000. The injection time was fixed to 150 ms resulting in a scan rate of 3.8 pixel/s, and the spatial resolution was 65 µm. A home-built Swagelok DESI sprayer was operated with a mixture of 95% methanol, 5% water delivered with a flow rate of 1.5 µL/min and nebulized with nitrogen at a backpressure of 6 bar. The resulting .raw files were converted into .mzML files using ProteoWizard msConvert (V.3.0.4043)(38) and subsequently compiled to an .imzML file (imzML converter V.1.3)(39). Metabolite identities were assigned based on comparing the observed mass to the theoretical mass for each metabolite (Suppl. Table 8). The mass difference between theoretical and observed was less than 2 ppm for all metabolites.

For image analysis, the ROIs were manually annotated on H&E images in QuPath(40), which were transferred to the corresponding MSI images using landmark-based two-dimensional affine transform on sample-by-sample basis. Relative abundances of metabolites were determined as mean intensity over pixels inside the masks. For a given mass-to-charge ratio, *R*, ion images were extracted by summing all intensities in the mass spectrum at each pixel on the interval *R* ± Δ*R*, where *ΔR* = 7×10^−6^ × *R*; intensities shown in heat maps in Fig. 5 were clipped at the 99^th^ percentile.

### Statistical analysis

Statistical analysis was performed in GraphPad Prism v.10.0.2 (Dotmatics, Boston, USA). Mean and standard deviation (SD) were used for normally distributed data, while median and interquartile range (IQR) summarised the non-parametric data, as probed with Shapiro-Wilk test for normality.

## Results

### Clinical features

An overview of the patient case is shown in Fig. 1. A male patient in his 70s underwent an abdominal ultrasound scan in February 2021 during which a 6.6 cm right lower-pole renal mass was discovered incidentally. Further characterisation on contrast-enhanced CT (ce-CT) revealed an isodense, solid, and predominantly endophytic mass, without regional or distant metastases (Fig. 1b). CT-guided biopsy revealed a high-grade papillary architecture with prominent eosinophilic nucleoli and perinucleolar clearing. Subsequent immunohistochemistry (IHC) showed loss of FH and a strongly positive S-(2-succino) cysteine (2SC) staining pattern (Fig. 1c). These findings were compatible with a diagnosis of FHd-RCC, and due to the potential aggressive behaviour of this subtype, radical nephrectomy was recommended and performed. The patient was subsequently referred to the clinical genetics team for germline genetic testing for a suspected HLRCC syndrome; there was no family history noted as the patient was adopted. No pathogenic variant or copy number changes of the *FH* gene were identified in the blood samples.

**Figure 1:**
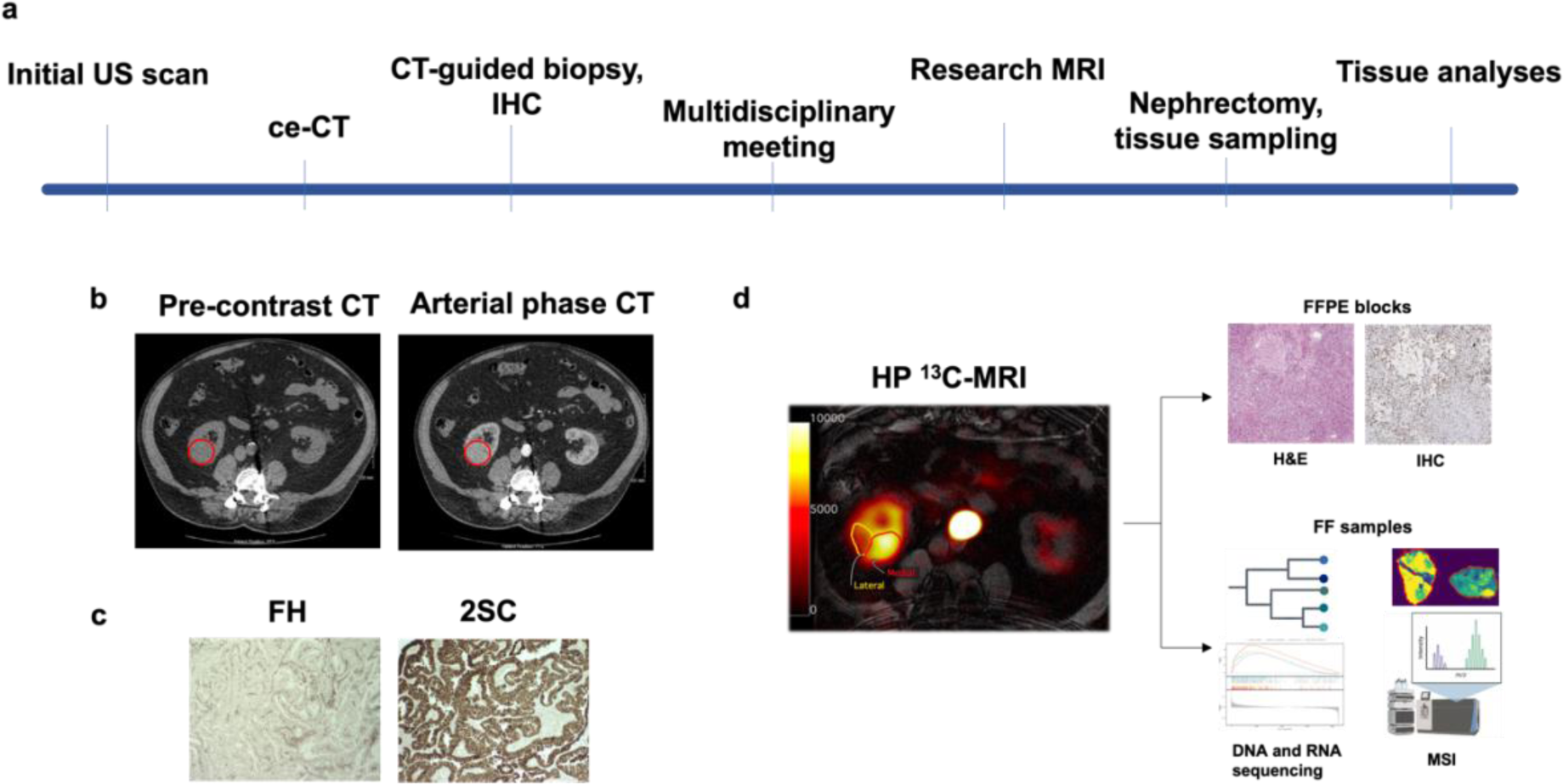
Overview of the case. (a) Timeline of the diagnostic work-up and research procedures undertaken on the patient. (b) Axial CT images pre-contrast and following contrast medium in the arterial phase demonstrating a right-sided isodense renal mass. (c) IHC staining of the CT-guided biopsy sample showed FH loss and strong 2SC positivity, consistent with a diagnosis of FHd-RCC. (d) Schematic representation of the research workflow with presurgical HP ^13^C-MRI imaging detecting two distinct metabolic tumour regions. Based on this difference, tissues were sampled post-nephrectomy, with subsequent analyses including H&E, IHC, DNAseq, RNAseq and MSI. US: ultrasound, CT: contrast-enhanced CT, IHC: immunohistochemistry, FH: fumarate hydratase, 2SC: S-(2-succino) cysteine, HP ^13^C-MRI: hyperpolarised [1-^13^C]pyruvate MRI, FFPE: formalin-fixed paraffin-embedded, FF: flash-frozen, MSI: mass spectrometry imaging.

Four days prior to the date planned for surgery, the patient underwent imaging with HP ^13^C-MRI, multiparametric ^1^H-MRI, including the dynamic contrast-enhanced (DCE)-MRI, diffusion-weighted imaging (DWI), and ^1^H-MRS as described in Methods. Based on the demarcation of two distinct regions on HP ^13^C-MRI signal within the tumour (Fig. 2), biopsies were collected from these tumour portions, as well as from adjacent normal-appearing kidney for further genomic, transcriptomic, proteomic and metabolomic analyses.

**Figure 2:**
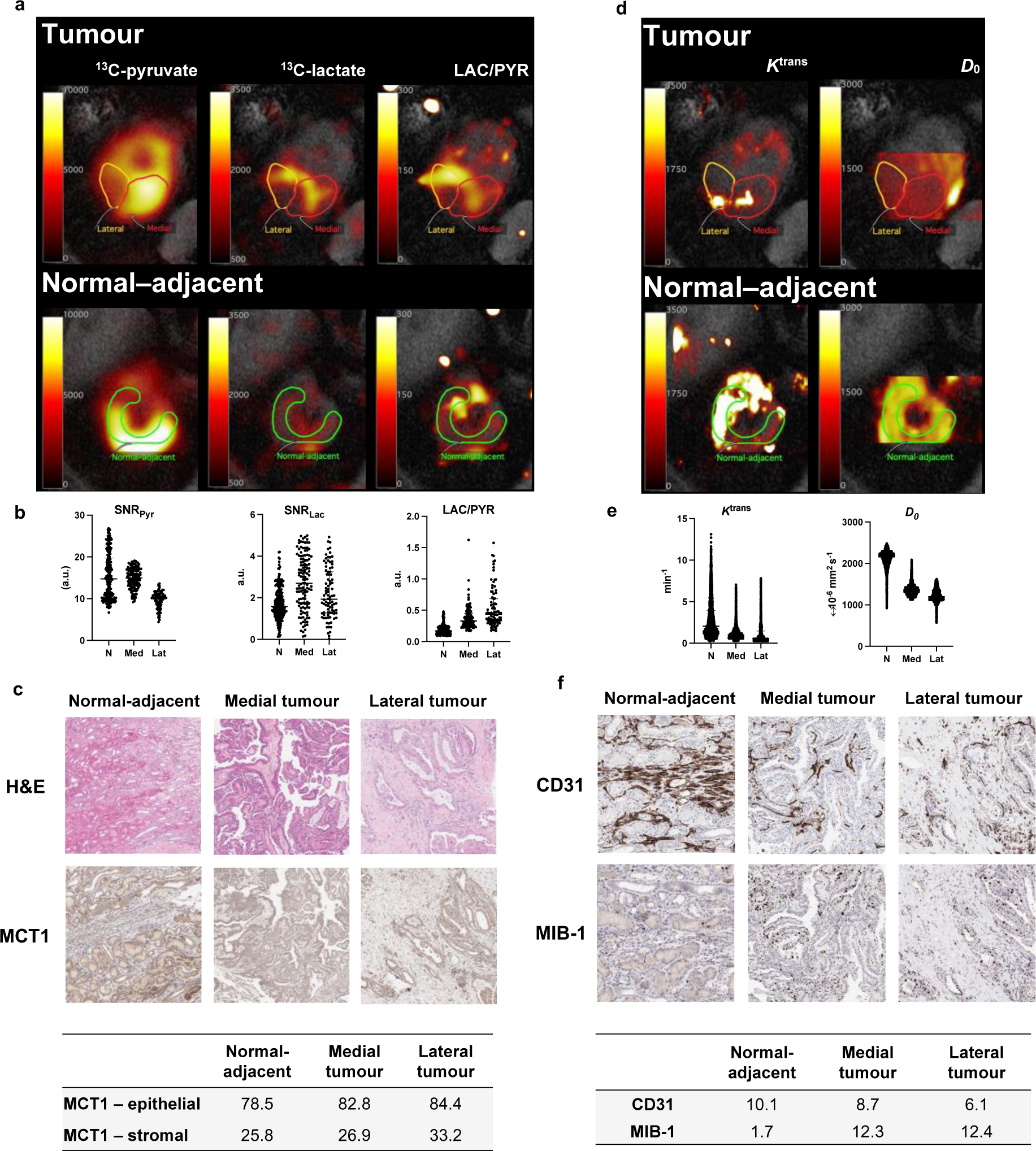
HP ^13^C-MRI from the FHd-RCC patient detects intratumoral metabolic heterogeneity as shown by the HP ^13^C-pyruvate-to-lactate conversion, while the normal adjacent kidney exhibits the highest perfusion and lowest cellularity. (a) HP ^13^C-MRI maps including summed ^13^C-pyruvate, summed ^13^C-lactate, and LAC/PYR are overlaid on axial T_1_ weighted (T_1_w) imaging. The top row shows the two metabolically distinct portions of the tumour with the normal-adjacent kidney delineated below. (b) The plots of voxel intensities of the HP ^13^C-MRI parameters depict the highest SNR_Pyr_ in the normal-adjacent kidney, and highest LAC/PYR ratio in the lateral tumour part. Medians are presented as black lines within individual values. The H&E and MCT1-stained images from the three different regions identified on HP ^13^C-MRI are shown in (c); MCT1 stained most strongly in the epithelial compartment of both tumour regions. (d) ^1^H-MRI-derived biomarker maps for perfusion and vascular leakage (*K*^trans^) and cellularity (*D*_0_), with the delineated ROIs, are shown. (e) Voxelwise quantification of these two biomarkers detected the highest perfusion in the normal-adjacent kidney with the least diffusion restriction, while the opposite was true for the lateral tumour region. (f) Representative IHC images of CD31 and MIB-1, with corresponding quantifications shown below. The high perfusion in the normal-adjacent kidney and confirmed by the strongest CD31 stain of endothelial cells here. MIB-1 as a proliferation marker was high in both tumour parts. IHC quantifications are presented as % positive cells. N = ipsilateral normal-appearing kidney, Med = medial portion of the tumour, Lat = lateral portion of the tumour.

The post-surgical evaluation confirmed the FHd-RCC subtype, and classified it as pT3a pN0 cM0 stage, with invasion of the renal vein but clear surgical margins.

### HP ^13^C-MRI detects metabolic heterogeneity by identifying a metabolically active intratumoral region

Metabolic maps of the ^13^C-pyruvate and ^13^C-lactate signals in each voxel, summed over the whole time-course, and their ratio LAC/PYR, are shown in Fig. 2a. Comparison of the medians of frequency distribution for each of the analysed regions-of-interest (ROIs) are plotted in Fig. 2b, with detailed statistical results in Suppl. Table 1. H&E and IHC stains for MCT1 (monocarboxylate transporter 1 which imports pyruvate) were analysed for each of the three ROIs and are shown in Fig. 2c, with quantification of the stains in the table below.

The same ROIs as those used for the HP ^13^C-MRI maps were transferred to co-registered quantitative maps derived from multiparametric ^1^H-MRI: *K*^trans^ measuring perfusion and vascular leakage of the contrast agent using DCE-MRI; and *D*_0_, the diffusion coefficient from DWI and an inverse measure of cellularity(41); Fig. 2d. Plots of individual voxel distributions are shown in Fig. 2e. IHC images of CD31 (a marker of endothelial cells and surrogate for microvascular density) and MIB-1 (a marker of cellular proliferation), with quantifications from the three regions as shown in Fig. 2f.

The signal-to-noise ratio for the hyperpolarised ^13^C-pyruvuate (SNR_Pyr_) was highest in the adjacent normal-appearing kidney (14.7) compared to both tumour regions (medial: 15.0; lateral: 10.2). However, conversion of ^13^C-pyruvate to ^13^C-lactate was lowest in the normal kidney compared to both portions of the tumour, as observed by the SNR_Lac_ (signal-to-noise ratio for the hyperpolarised ^13^C-lactate; normal kidney: 1.6; medial tumour region: 2.7; and lateral tumour region: 1.9), as well as by the LAC/PYR ratio (^13^C-lactate-to-pyruvuate ratio; normal kidney: 0.17; medial tumour region: 0.33; lateral tumour region: 0.44). Expression of the MCT1 transporter has previously been shown to relate to the hyperpolarised ^13^C-lactate signal and tumour aggressiveness in *FH* wild type ccRCC(20,42), but in this FHd-RCC its expression was similar in the tumour regions compared to the normal kidney (82.8% and 84.4% vs. 78.5%).

*K*^trans^ analysis showed the highest perfusion and vascular leakiness in the normal-adjacent kidney (2.0 min^−1^) compared to both tumour regions (medial: 1.0 min^−1^; lateral: 0.6 min^−1^); CD31 staining was highest in the normal tissue with 10% positive cells, compared to 6.1% and 8.7% in the tumour. Diffusion measurements using the diffusion coefficient (*D*_0_) was highest in the normal-adjacent kidney (2.16×10^−3^ mm^2^ s^−1^), with lower values in both tumour regions in keeping with increased cell density (medial: 1.35×10^−3^ mm^2^ s^−1^; lateral: 1.20×10^−3^ mm^2^ s^−1^). This was supported by the expression of the proliferation marker MIB-1 which was higher in both tumour regions compared to the adjacent normal kidney (normal kidney: 1.7%; medial tumour region: 12.4%; lateral tumour region: 12.3%).

^1^H-MRS acquisition was classified as a technical failure due to the unreliable detection of the choline peak due to baseline distortion and broad peaks(43) (Suppl. Fig. 1), which was due to the presence of spinal laminectomy screws close to the tumour (seen as beam hardening artefacts on CT image in Fig. 1) likely to have resulted in inhomogeneities in the local magnetic field(14).

### Genotyping reveals a germline FH variant previously undetected in this patient and confirms differential somatic FH loss across the tumour

Although routine clinical genetic testing showed no pathogenic variant or copy number variations (CNVs) in the venous blood sample, next generation sequencing (NGS) of postoperative tissue samples detected a *FH* variant of uncertain significance (VUS) in the normal-adjacent kidney sample (Fig. 3). The variant was an intronic indel c.1237-50TC, previously reported in HLRCC(44) but there is insufficient evidence of the pathogenicity of this variant.

**Figure 3:**
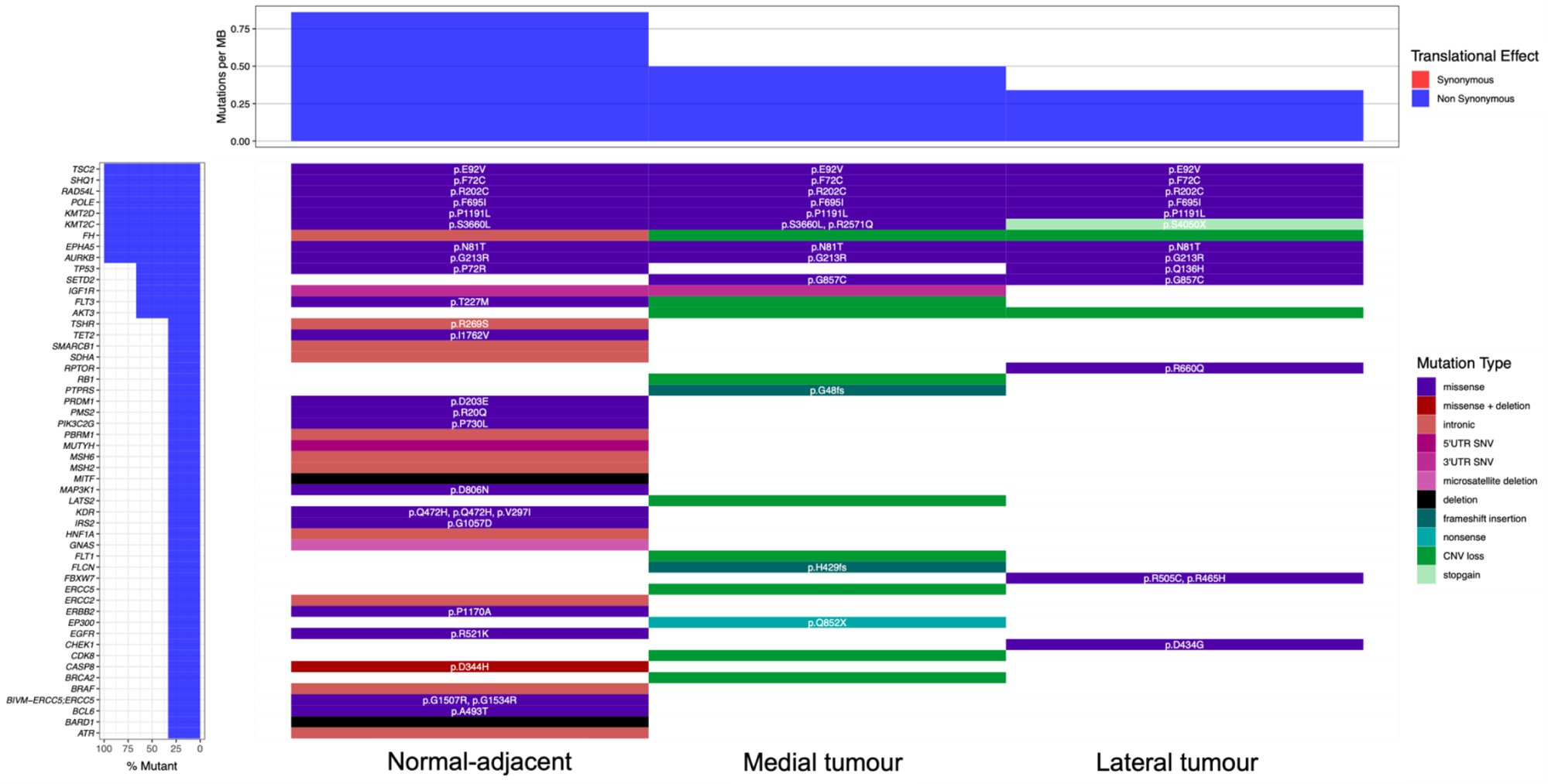
Genetic analysis of the three samples of FHd-RCC case. Waterfall plot of genetic aberrations found across the samples shows a *FH* mutation in the germline sample and CNV loss in both tumour parts. Eight further mutations were identified across all samples, including *TSC2*, *SHQ1*, *RAD54L*, *POLE*, *KMT2D*, *KMT2C*, *EPHA5* and *AURKB*. Divergence of mutations was detected between the two tumour portions suggesting intratumoral genetic heterogeneity, with the lateral tumour portion carrying confirmed oncogenic mutations in *FBXW7*. CNV: copy number variation; SNV: single nucleotide variant.

However, the medial and lateral tumour portions harboured somatic aberrations in *FH*, including the copy number variation (CNV) showing a −2.24 log fold change in the medial tumour region, and a −0.78 log fold change in the lateral portion, therefore confirming the *FH* loss on a genetic level. Genomic variations of the three samples are summarised in the waterfall plot in Fig. 3, which indicates eight further mutations identified across all samples, including: *TSC2*, *SHQ1*, *RAD54L*, *POLE*, *KMT2D*, *KMT2C*, *EPHA5* and *AURKB*. Of these, only *RAD54L* c604C>T:p.R202C is classified as likely to be pathogenic in both germline and somatic contexts(45), while *SHQ1* p.F72C, *EPHA5* p.N81T, and *AURKB* p.G213R are classified as VUS, scoring above 0.60 (for further details on the score algorithm, refer to Methods), therefore raising the possibility of pathogenicity. An additional variant in the normal-adjacent sample was detected which is of potential pathogenic clinical significance but this was not found in the tumour samples: a missense mutation in *KDR* c.1416A>T:p.Q472H(46). Interestingly, comparison between the two tumour samples showed a degree of genetic divergence with pathogenic variants detected in the medial tumour part showing a frameshift mutation in *FLCN* p.H429fs and an exonic stopgain in *EP300* p.Q825X. However, the lateral tumour region harboured mutations in *FBXW7* which are known to be oncogenic (p.R505C) or likely to be oncogenic (p.R465H), as classified by the OncoKB database(47). Overlapping mutations between both the medial and lateral parts were as follows: a pathogenic stop gain mutation in *KMT2C* p.S4050X and missense substitutions with a score > 0.60 in TP53 p.Q136H, *RPTOR* p.R660Q, and *SETD2* p.G875C.

### Mass Spectrometry Imaging confirms metabolic compartmentalisation within the tumour and across cellular compartments

FHd cells accumulate fumarate due to the inability to convert it to downstream malate, leading to a block in the TCA cycle with a subsequent shift towards enhanced aerobic glycolysis to meet energetic requirements(10). A more detailed metabolic analysis of the tumour was undertaken using Desorption Electrospray Ionisation (DESI) Mass Spectrometry Imaging (MSI); maps of the detected metabolites of interest with their relative quantitation within the three regions are shown in Fig. 4.

**Figure 4:**
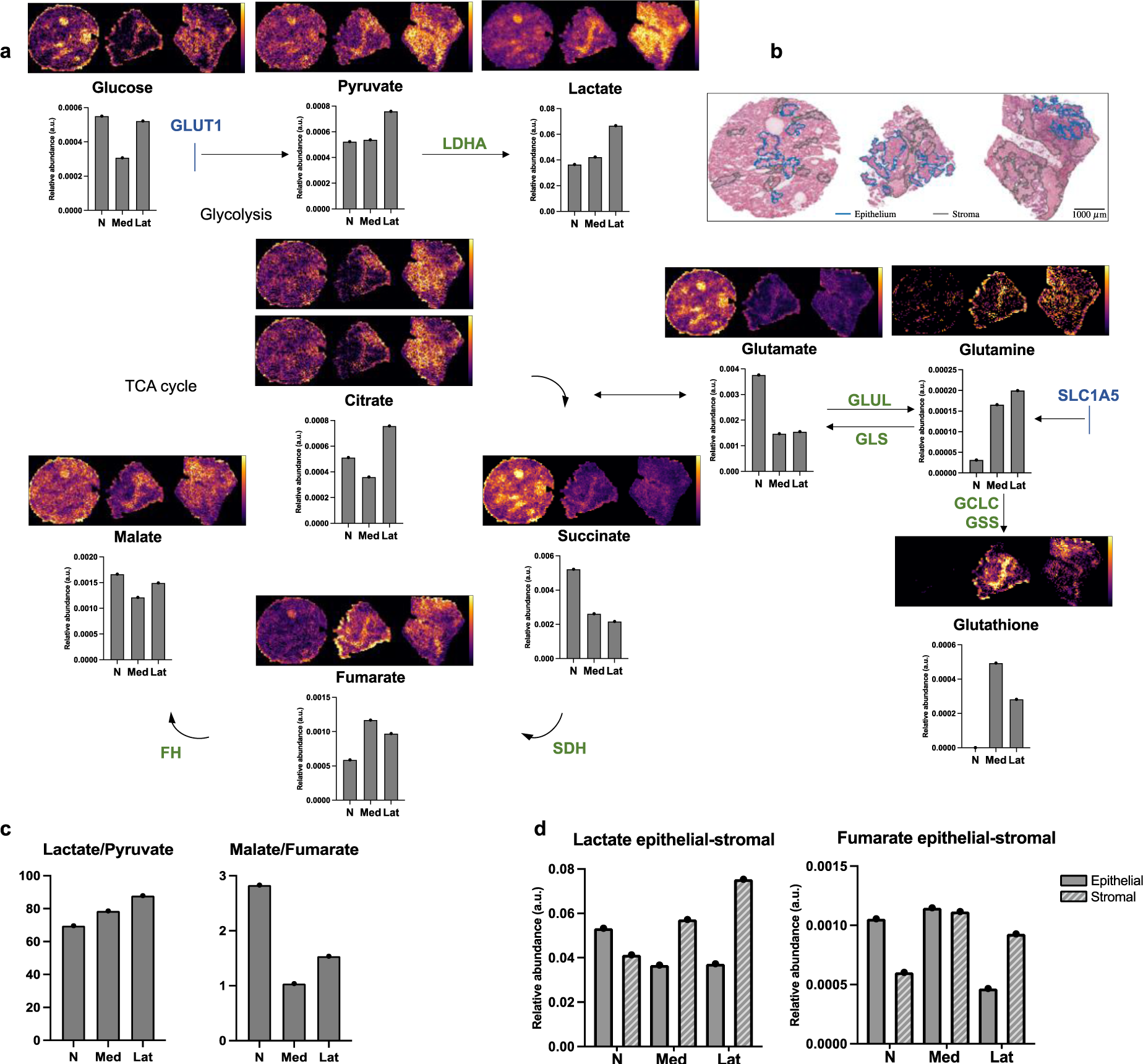
MSI identifies increased lactate generation and decreased fumarate-to-malate conversion in the tumour compared to normal tissue and confirms intratumoral heterogeneity observed on metabolic MRI and genotyping. (a) Diagram of relevant metabolites (relative abundance in arbitrary units plotted in barplots), transporters (highlighted in dark blue), and enzymes (highlighted in green) involved in metabolic pathways of interest, measured across the three regions of interest. Images show the distribution of the metabolites across the tissue section. (b) H&Es of the three samples with manually annotated regions of interest (blue = epithelial; grey = stromal) which were transferred to MSI images for analysis of the metabolic compartmentalisation. (c) The lactate/pyruvate and malate/fumarate ratios were determined within the three regions suggesting the highest pyruvate-to-lactate conversion in the lateral tumour region and the lowest in the normal-adjacent kidney. The opposite trend was found for malate-to-fumarate conversion with the lateral region exhibiting a higher malate/fumarate ratio compared with the medial tumour part, confirmed by higher *FH* expression on RNAseq. (d) Barplots presenting lactate and fumarate quantified per ROIs. N: normal-adjacent kidney, Med: medial tumour region, Lat: lateral tumour region, GLUT1: glucose transporter 1, LDHA: lactate dehydrogenase A, MPC: mitochondrial pyruvate carrier, PDH: pyruvate dehydrogenase, GLUD1: glutamate dehydrogenase, GLUL: glutamine synthetase, GLS: glutaminase, GCLC: glutamate-cysteine ligase, GSS: glutathione synthetase, SDH: succinate dehydrogenase, FH: fumarate dehydrogenase.

As expected, the fumarate concentration was 1.6 to 2.0-times higher in the tumour compared to the normal kidney (normal kidney: 5.9 × 10^−4^ a.u.; medial tumour: 11.7 × 10^−4^ a.u.; lateral tumour: 9.7 × 10^−4^ a.u.), and the malate/fumarate ratio was correspondingly 1.9 to 2.8-times lower, confirming the low conversion of fumarate to malate via FH (malate/fumarate ratio: 2.8 in normal kidney; 1.0 in the medial tumour; 1.5 in the lateral tumour). The higher accumulation of fumarate in the medial portion of the tumour compared to the lateral part, correlated with a lower expression of *FH* on both RNA sequencing and on genotyping (medial tumour: 67 counts; lateral tumour: 192 counts; Suppl. Table 4).

The fumarate distribution across cell compartments was lost in the tumour samples compared to the normal kidney, where fumarate is normally contained in epithelium and is actively reabsorbed from the lumen and from the interstitium via the sodium-dicarboxylate cotransporters 1 (NaDC1) and 3 (NaDC3), respectively(48). This cotransporter expression was profoundly decreased in these tumour samples which showed very low *NaDC1* and *NaDC3* RNAseq counts (Suppl. Table 4). Moreover, the relative fumarate compartmentalisation between the stroma and epithelium was also altered with tumour formation, with an equal distribution of fumarate in the epithelial and stromal compartments in the medial tumour, and a reversal of the fumarate distribution in the lateral portion (Fig. 4d).

The differences in ^13^C-pyruvate-to-lactate conversion on HP ^13^C-MRI between the tumour regions was supported by the lactate/pyruvate ratio on MSI which was 1.1 to 1.3-times higher in the tumour compared to normal (lactate/pyruvate ratio: 69.6 in normal kidney; 78.7 in the medial tumour; 87.8 in the lateral tumour). The normalised RNAseq counts for *LDHA,* one of the subunits of the enzyme LDH, were also higher in the tumour compared to normal (normal: 41,936 counts; medial tumour: 61,931 counts; lateral tumour: 131,521 counts; Suppl. Table 4). The lactate concentration was higher in the stroma compared to the epithelium, with a stromal-to-epithelial lactate ratio of 1.5 to 2.0. This may be partly explained by the MCT4 expression, as a major exporter of lactate, which showed higher expression levels on the tumour epithelial cells (positive cell counts; epithelium: 77% and 94% vs. stroma: 62% and 49%, in medial and lateral tumour regions respectively; Suppl. Table 5).

Interestingly, despite the known TCA cycle disruption in the FHd cells, the lateral tumour portion still exhibited a 1.5-times higher concentration of citrate compared to the normal tissue (normal kidney: 5.1 × 10^−4^ a.u.; medial tumour: 3.6 × 10^−4^ a.u.; lateral tumour: 7.6 × 10^−4^ a.u.). This finding was supported by the expression of citrate synthase (*CS*), which converts acetyl-CoA to citrate, which was also highest in the lateral tumour portion (normal kidney: 9,373 counts; medial tumour: 7,041 counts; lateral tumour: 13,432 counts; Suppl. Table 4). However, the intratumoral succinate levels were 0.4 to 0.5-times lower than in the normal adjacent kidney (normal kidney: 5.2 × 10^−3^ a.u.; medial tumour: 2.6 × 10^−3^ a.u.; lateral tumour: 2.2 × 10^−3^ a.u.), despite the normal RNA expression of *SDHA*, a subunit of SDH which catalyses the conversion of succinate to fumarate (normal kidney: 2,505 counts; medial tumour: 2,393 counts; lateral tumour: 3,095 counts; Suppl. Table 4). This finding may reflect upstream shunting of α-ketoglutarate to glutamine, and subsequently glutathione, for protection against oxidative damage secondary to the downstream impairment in the TCA cycle(10). To support this hypothesis, glutathione was shown to be 486 to 851-times higher in the tumour compared to normal (normal kidney: 0.6 × 10^−6^ a.u.; medial tumour: 493.8 × 10^−6^ a.u.; lateral tumour: 282.0 × 10^−6^ a.u.). In addition, the elevated tumour glutamine (normal kidney: 0.3 × 10^−4^ a.u.; medial tumour: 1.7 × 10^−4^ a.u.; lateral tumour: 2.0 × 10^−4^ a.u.) was likely to be derived from intracellular metabolism, as there was a lower expression of the neutral amino acid transporter, *SLC1A5*, which facilitates glutamine membrane transport (normal kidney: 12,282 counts; medial tumour: 712 counts; lateral tumour: 551 counts; Suppl. Table 4).

### FHd-RCC exhibited high immunogenicity with CD8+ T cell infiltration in the most metabolically active tumour region

Finally, we investigated the cell-type composition of each sample, to understand the underlying cellular drivers for the changes in metabolism as this may have implications for therapeutic approaches in this rare tumour type, which is currently treated with antiangiogenic therapy(4). Using RNAseq deconvolution with BayesPrism(35) and CD8 staining, we investigated the differences in the cellular composition between the three samples (Fig. 5).

**Figure 5:**
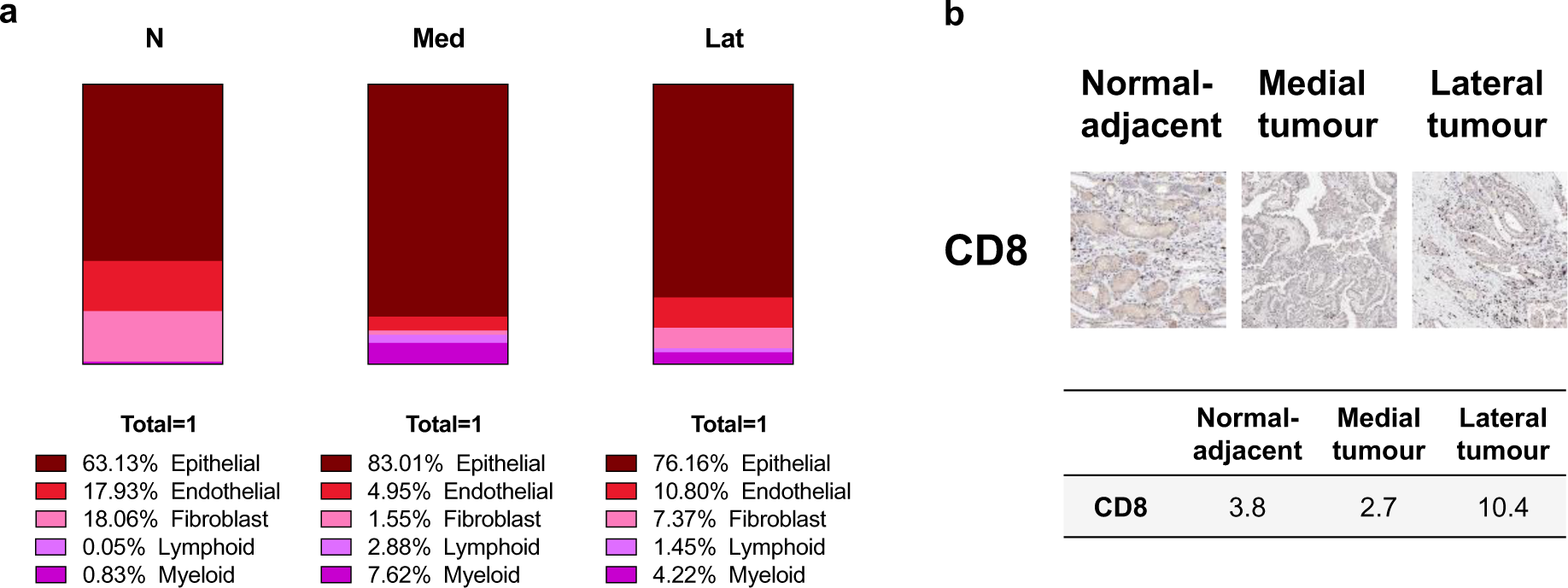
RNAseq deconvolution and CD8 staining of the three different regions of interest. (a) Fractional cellular composition within each sample, suggesting lower endothelial and higher immune cell compartments (lymphoid and myeloid) in the tumour compared to normal. N: normal-adjacent kidney, Med: medial tumour part, N: lateral tumour part. (b) The percentage of CD8+ T cells was highest in the lateral tumour part. IHC quantifications are presented as % positive cells.

Fig. 5a demonstrates that the endothelial compartment fraction in both tumour samples was lower compared to the normal sample (normal: 17.9%; medial tumour: 5.0%; lateral tumour: 10.8%), correlating with the perfusion imaging on ^1^H-MRI, which identified lower perfusion in the tumour compared to the normal-adjacent kidney (Fig. 2). The tumour samples contained a much higher proportion of immune cells compared to the normal samples: the lymphoid proportion was 29 to 58-times higher in the tumour compared to the normal tissue. Myeloid infiltration was also increased in the tumour, although to a lesser degree (5 to 9-times higher).

Interestingly, the IHC staining for CD8+ T cells was increased only in the lateral tumour part (10.4% positive cells) compared to the normal-adjacent kidney (3.8%), while the medial tumour part stained the lowest (2.7% positive cells), as shown in Fig. 5b. These findings are in line with previous reports of high immunogenicity within FHd-RCC(49).

## Discussion

FHd-RCC is a highly aggressive kidney cancer subtype but is challenging to manage due to its rarity, which hinders the development of evidence-based clinical guidelines for treatment. While the molecular effects of FH-loss have been extensively studied, intratumoral heterogeneity exists across many scales *in vivo* and is difficult to evaluate using invasive approaches. Proton magnetic resonance spectroscopy (^1^H-MRS) has been used to detect high levels of fumarate in patients with FH loss but has limited spatial resolution, is technically not possible in all patients, and cannot probe many other altered metabolites found at lower concentration(14). The development of novel metabolic imaging tests for these tumours will aid accurate diagnosis and tumour characterisation and patient prioritisation for appropriate treatment. Non-invasive imaging can be used to probe structural, functional, and cellular heterogeneity within tumours, and here we have demonstrated how it can be applied to study tumour metabolism in a case of FHd-RCC. This report has demonstrated intratumoral metabolic heterogeneity, both macroscopically and microscopically, using novel methods such as metabolic MRI and mass spectrometry imaging of tissue samples.

The FHd-RCC tumour exhibited relatively poor perfusion compared to the normal kidney, which renders the assessment of aggressiveness on routine clinical contrast enhanced CT alone difficult. Metabolic MRI with hyperpolarised ^13^C-pyruvate demonstrated the production of labelled tissue lactate, a parameter known to correlate with tumour aggressiveness in other tumours including clear cell renal cell carcinoma(20,21). A higher LAC/PYR ratio was observed in the tumour compared to the normal kidney, and intratumoral metabolic heterogeneity was also demonstrated prompting subsequent multiregional tissue analyses to discern the drivers for the observed spatial variation in metabolism.

Although the tumour region with the higher LAC/PYR ratio harboured a smaller log fold reduction in *FH* expression in comparison to the more medial tumour, it carried other mutations recognised as oncogenic(47). The relative preservation of *FH* may represent dedifferentiation because of the acquisition of these additional oncogenic mutations during tumour progression. An analogy of this is found in high grade *FH*-wildtype ccRCC which is known to undergo dedifferentiation from the initial Von Hippel-Lindau (VHL) related tumorigenesis(50). In addition, a VUS of *FH* was detected in the normal-adjacent kidney, therefore raising the possibility of a diagnosis of HLRCC in this patient given the clinical presentation of renal cell cancer. This VUS has been reported in the context of this syndrome previously(44), but there is insufficient evidence of its pathogenicity.

The metabolic intratumoural heterogeneity identified on imaging was validated on the tissue level using DESI Mass Spectrometry Imaging. A high lactate/pyruvate ratio on MSI was observed in the tumour region with the higher LAC/PYR ratio on HP ^13^C-MRI. Furthermore, the divergent genetic *FH*-loss was confirmed by increased fumarate accumulation and a lower fumarate-to-malate conversion in the tumour region with greater *FH*-loss. There was an inverse relationship between lactate concentration and fumarate accumulation on MSI within the tumour and this mismatch between the lactate concentration and fumarate accumulation between regions could be a result of additional oncogenic mutations as detected on sequencing. This result also emphasises the importance of measuring multiple metabolites to fully characterise the metabolic tumour phenotype and potentially this could help in the assessment of the underlying tumour aggressiveness. The high fumarate concentration in FHd cells potentiates tumorigenic activity, and in tandem with additional mutations *in vivo*, can result in enhanced tumour aggressiveness(51). Regional pH may also play a role as the lactate rich region is likely to be acidic, and fumarate has been shown to have increased succination-activity in a low pH environment which may contribute to the oncogenic effect(52).

These results provide a rationale for using HP ^13^C-MRI to characterise tumour metabolism *in vivo* and to guide biopsies to areas of greatest metabolic importance. Although ^1^H-MRS can be used to detect endogenous fumarate accumulation directly, sensitive detection of lactate can be challenging, and HP ^13^C-MRI can also detect dynamic metabolism. ^1^H-MRS may also be prone to technical failure as shown here, from susceptibility to magnetic field inhomogeneities and long acquisition times resulting in motion artefacts(15). In comparison, HP ^13^C-MRI acquisition is very rapid and can be undertaken in a single breath hold to provide information on both pyruvate delivery as a measure of perfusion, and metabolic conversion to lactate. The importance of evaluating both perfusion and metabolism has previously been recognised on FDG-PET studies in other tumour types(53–55), and perfusion-metabolism mismatch is a recognised feature of aggressive tumours outgrowing their blood supply(56). Furthermore, given the inverse relationship between lactate and fumarate on MSI within the tumour, ^1^H-MRS and HP ^13^C-MRI could be used to target high levels of fumarate and lactate within the tumour respectively, revealing complementary metabolic information enabling the tumour phenotype and its heterogeneity to more accurately studied.

Finally, an increased immune fraction was demonstrated in both tumour regions compared to the normal-adjacent kidney, with a very high infiltration of CD8+ T cells in the lactate-rich area. However, despite the presence of large numbers of T cells, their antitumour function may be suppressed due to the hostile microenvironmental conditions in the FHd tumour, including high lactate generation and fumarate accumulation, which have previously been reported to impair T cell activity and cytokine production(57,58). Furthermore, a suppressed immune microenvironment has been reported in SDH-deficient phaeochromocytomas and paragangliomas(59,60). This may have important clinical considerations given that FHd-RCC is managed as a “non-clear cell RCC” under the European Society for Molecular Oncology (ESMO) guidelines, which recommend first-line antiangiogenic therapy(4). However, in the context of relatively low perfusion/vascular leakage and significant immune infiltration as shown in this FHd-RCC, the patient may benefit from immunotherapy as suggested by previous reports(49,61,62). Future research into therapeutic approaches for FHd-RCC could investigate the effect of combining immunotherapy with metabolic modulation of the TME(63).

The implications of studying intratumoral metabolic heterogeneity and compartmentalisation are far-reaching. Metabolic imaging could be used to target a biopsy to the most metabolically active tumour region, which in turn could provide appropriate samples for accurate metabolic phenotyping on a tissue level. This approach of metabolic imaging across scales could be used to improve diagnosis, characterisation, and treatment planning. Furthermore, understanding metabolic heterogeneity, and combining this with other features of the TME such as perfusion and diffusion, could be used to select and monitor the effects of conventional therapeutic agents based on evaluating the most metabolically active region. These techniques could also be used to develop novel metabolic therapeutics targeted to either the tumour or the immune compartments. Lastly, HP ^13^C-MRI could be used to detect small yet highly aggressive and metabolically active tumours, which could lead to earlier and more effective treatment. This report has demonstrated the potential of imaging tumour metabolism across multiple scales using FHd-RCC as a model system.

## Acknowledgements

Authors acknowledge funders and the technical support from the radiographers’ team of the MRI and Spectroscopy Unit at Addenbrooke’s Hospital, Cambridge University Hospitals.

## Authors’ contributions

Conceived and designed the experiments: I.H.M., R.C., C.Y., J.K., J.W., J.D., A.Y.W., S.J.W., T.A., T.J.M., G.D.S., M.A.M., F.A.G. Performed the experiments: I.H.M., G.H., H.H., A.S.K., J.K., A.B.G., A.N.P., C.B., S.A., M.D., K.H. Analysed the data: I.H.M., J.D., G.H., H.H., J.G., A.S.K., J.K., A.B.G., A.N.P., C.B., S.A., M.D., K.H., T.J.M. Wrote the main paper: I.H.M., F.A.G. Review and edits of the paper: all co-authors. All authors have read and agreed to the published version of the manuscript.

## Ethics approval and consent to participate

This study was performed in accordance with Declaration of Helsinki. The participant gave written consent for ethically approved studies MISSION Substudy in Renal Cancer (Research Ethics Committee (REC) number: 15/EE/0378), ARTIST (REC: 20/EE/0200) and EMKC (REC: 19/EE/0161).

## Consent for publication

Not applicable.

## Data availability

All data that support the findings of this study are available on reasonable request from the corresponding author, on condition that this will not be used to deanonymize the patient. The data are not publicly available due to them containing information that could compromise research participant privacy and consent.

