## Supplementary for "Probing intratumoral metabolic compartmentalisation in fumarate hydratase-deficient renal cancer using clinical hyperpolarised ^13^C-MRI and mass spectrometry imaging"

| | **$SNR_{Pyr}$** [a.u.] | **$SNR_{Lac}$** [a.u.] | **LAC/PYR** [a.u.] |
|---|---|---|---|
| **Normal-adjacent** | 14.7 (10.2-19.8) | 1.6 (1.2-2.3) | 0.17 (0.13-0.23) |
| **Medial tumour part** | 15.0 (13.0-16.9) | 2.7 (1.7-3.9) | 0.33 (0.25-0.45) |
| **Lateral tumour part** | 10.2 (8.5-11.1) | 1.9 (1.3-3.2) | 0.44 (0.31-0.68) |

**Supplementary Table 1: Statistical results of comparison of voxelwise intensities of the HP $^{13}$C-MRI parameters of the FHd-RCC case.** All results are presented as median ± IQR.

| | $K^{trans}$ [min$^{-1}$] | $D_0$ [× 10$^{-3}$ mm$^2$ s$^{-1}$] |
|---|---|---|
| **Normal-adjacent** | 2.05 (1.21-3.96) | 2.16 (2.03-2.24) |
| **Medial tumour part** | 1.00 (0.72-1.49) | 1.35 (1.27-1.42) |
| **Lateral tumour part** | 0.57 (0.39-1.48) | 1.20 (1.13-1.33) |

**Supplementary Table 2: Statistical results of comparison of voxelwise intensities of the $^1$H imaging biomarkers of the FHd-RCC case.** All results are presented as median ± IQR.

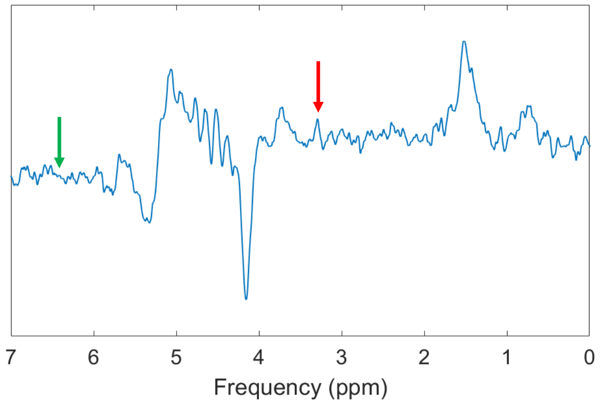


**Supplementary Figure 1: $^1$H-MRS spectrum.** MRS spectrum: the red arrow shows the expected location for the choline peak at 3.2 ppm, and the green arrow the expected location of the fumarate peak at 6.54 ppm. Baseline distortion, broad peaks (FWHM = 38 Hz) and an unreliable choline peak classified the $^1$H-MRS acquisition as a technical failure.

| | *LDHA* | *MPC1* | *PDHA1* | *CS* | *GLUL* | *GLS* | *GCLC* | *GSS* | *SLC1A5* | *SDHA* | *FH* | *NaDC1* | *NaDC3* |
|---|---|---|---|---|---|---|---|---|---|---|---|---|---|
| **Normal-adjacent** | 41936 | 5406 | 9273 | 9373 | 19742 | 34683 | 2245 | 5026 | 12282 | 2505 | 6118 | 5357 | 39643 |
| **Medial tumour** | 61932 | 429 | 5019 | 7041 | 9280 | 12153 | 1815 | 5556 | 712 | 2393 | 67 | 6 | 40 |
| **Lateral tumour** | 131521 | 1949 | 8996 | 13432 | 19163 | 20237 | 8460 | 5594 | 551 | 3095 | 192 | 3 | 17 |

**Supplementary Table 3: Normalised RNAseq counts for the enzymes relevant to metabolic conversions of interest.** LDHA = lactate dehydrogenase A, PDH = pyruvate dehydrogenase, SDH = succinate dehydrogenase, FH = fumarate dehydrogenase.

| | MCT4 epithelial | MCT4 stromal |
|---|---|---|
| **Normal-adjacent** | 63 | 30 |
| **Medial tumour** | 77 | 62 |
| **Lateral tumour** | 94 | 49 |

**Supplementary Table 4: Quantification of MCT4 stain on IHC.** Results are presented as % positive cells. MCT4 = monocarboxylate transporter 4.

| | | | | | | |
|---|---|---|---|---|---|---|
| ABL1 | CD79B | ESR1 | IGF1 | KMT2C | PIM1 | SDHD |
| AKT1 | CDC73 | ETV1 | IGF1R | MPL | PLK2 | SETD2 |
| AKT2 | CDH1 | ETV6 | IGF2 | MRE11A | PMAIP1 | SF3B1 |
| AKT3 | CDK12 | EZH2 | IKBKE | MSH2 | PMS1 | SH2D1A |
| ALK | CDK4 | AMER1 | IKZF1 | MSH6 | PMS2 | SHQ1 |
| ALOX12B | CDK6 | FAM175A | IL10 | MTOR | PNRC1 | SMAD2 |
| APC | CDK8 | FAM46C | IL7R | MUTYH | POLD1 | SMAD3 |
| AR | CDKN1A | FANCA | INPP4A | MYC | POLE | SMAD4 |
| ARAF | CDKN1B | FANCC | INPP4B | MYCL | PPP2R1A | SMARCA4 |
| ARID1A | CDKN2A | FANCL | INSR | MYCN | PPP2R2A | SMARCB1 |
| ARID1B | CDKN2B | FANCM | IRAK4 | MYD88 | PRDM1 | SMARCD1 |
| ARID2 | CDKN2C | FAT1 | IRF4 | MYOD1 | PRKAR1A | SMO |
| ARID5B | CHEK1 | FBXW7 | IRS1 | NBN | PTCH1 | SOCS1 |
| ASXL1 | CHEK2 | FGF19 | IRS2 | NCOR1 | PTEN | SOX17 |
| ASXL2 | CIC | FGF3 | JAK1 | NF1 | PTPN11 | SOX2 |
| ATM | CREBBP | FGF4 | JAK2 | NF2 | PTPRD | SOX9 |
| ATR | CRKL | FGFR1 | JAK3 | NFE2L2 | PTPRS | SPEN |
| ATRX | CRLF2 | FGFR2 | JUN | NKX2-1 | PTPRT | SPOP |
| AURKA | CSF1R | FGFR3 | KDM5A | NKX3-1 | RAC1 | SRC |
| AURKB | CTCF | FGFR4 | KDM5C | NOTCH1 | RAD50 | STAG2 |
| AXIN1 | CTLA4 | FH | KDM6A | NOTCH2 | RAD51 | STK11 |
| AXIN2 | CTNNB1 | FLCN | KDR | NOTCH3 | RAD51B | STK40 |
| AXL | CUL3 | FLT1 | KEAP1 | NOTCH4 | RAD51C | SUFU |
| B2M | DAXX | FLT3 | KIT | NPM1 | RAD51D | SUZ12 |
| BAP1 | DCUN1D1 | FLT4 | KLF4 | NRAS | RAD52 | SYK |
| BARD1 | DDR2 | FOXA1 | KRAS | NSD1 | RAD54L | TBX3 |
| BBC3 | DICER1 | FOXL2 | LATS1 | NTRK1 | RAF1 | TERT |
| BCL2 | DIS3 | FOXP1 | LATS2 | NTRK2 | RARA | TET1 |
| BCL2L1 | DNMT1 | FRS2 | LMO1 | NTRK3 | RASA1 | TET2 |
| BCL2L11 | DNMT3A | FUBP1 | MAD2L2 | PAK1 | RB1 | TGFBR1 |
| BCL6 | DNMT3B | GATA1 | MALT1 | PAK7 | RBM10 | TGFBR2 |
| BCOR | DOT1L | GATA2 | MAP2K1 | PALB2 | RECQL4 | TMEM127 |
| BLM | E2F3 | GATA3 | MAP2K2 | PARK2 | REL | TMPRSS2 |
| BMPR1A | EED | GNA11 | MAP2K4 | PARP1 | RET | TNFAIP3 |
| BRAF | EGFL7 | GNAQ | MAP3K1 | PAX5 | RFWD2 | TNFRSF14 |
| BRCA1 | EGFR | GNAS | MAP3K13 | PBRM1 | RHEB | TOP1 |
| BRCA2 | EIF1AX | GREM1 | MAPK1 | PDCD1 | RHOA | TP53 |
| BRD4 | EP300 | GRIN2A | MAX | PDGFRA | RICTOR | TP63 |
| BRIP1 | EPCAM | GSK3B | MCL1 | PDGFRB | RIT1 | TRAF7 |
| BTK | EPHA3 | H3F3C | MDC1 | PDPK1 | RNF43 | TSC1 |
| CARD11 | EPHA5 | HGF | MDM2 | PHOX2B | ROS1 | TSC2 |
| CASP8 | EPHB1 | HIST1H1C | MDM4 | PIK3C2G | RPS6KA4 | TSHR |
| CBFB | ERBB2 | HIST1H2B D | MED12 | PIK3C3 | RPS6KB2 | U2AF1 |
| CBL | ERBB3 | HIST1H3B | MEF2B | PIK3CA | RPTOR | VHL |
| CCND1 | ERBB4 | HNF1A | MEN1 | PIK3CB | RUNX1 | VTCN1 |
| CCND2 | ERCC2 | HRAS | MET | PIK3CD | RYBP | WT1 |
| CCND3 | ERCC3 | ICOSLG | MITF | PIK3CG | SDHA | XIAP |
| CCNE1 | ERCC4 | IDH1 | MLH1 | PIK3R1 | SDHAF2 | XPO1 |
| CD274 | ERCC5 | IDH2 | KMT2A | PIK3R2 | SDHB | YAP1 |
| CD276 | ERG | IFNGR1 | KMT2D | PIK3R3 | SDHC | YES1 |

Supplementary Table 5: Custom gene list targeted for next generation sequencing.

| **IHC marker** | **Manufacturer** | **Catalogue Number** | **Host Species** |
|---|---|---|---|
| MCT1 (SLC16A1) | Atlas | HPA003324 | Rabbit |
| MCT4 (SLC16A3) | Atlas | HPA021451 | Rabbit |
| CD31 | Dako | M0823 | Mouse |
| MIB-1 | Dako | M7240 | Mouse |
| CD8 | Neomarkers | RM-9116-S | Rabbit |

Supplementary Table 6: Antibodies used for IHC staining.

| | Theoretical m/z | Observed m/z | ppm error |
|---|---|---|---|
| Glucose | 215.0328 [M+Cl]- | 215.03268 | -0.56 |
| Pyruvate | 87.0088 [M-H]- | 87.00865 | -1.72 |
| Lactate | 89.0244 [M-H]- | 89.0243 | -1.12 |
| Citrate | 191.0197 [M-H]- | 191.0198 | 0.52 |
| Succinate | 117.0193 [M-H]- | 117.01942 | 1.03 |
| Fumerate | 115.0037 [M-H]- | 115.00376 | 0.52 |
| Malate | 133.0142 [M-H]- | 133.0144 | 1.5 |
| Glutamate | 146.04588 [M-H]- | 146.046 | 0.82 |
| Glutamine | 145.06187 [M-H]- | 145.062 | 0.9 |
| Glutathione | 306.0765 [M-H]- | 306.0766 | 0.33 |

Supplementary Table 8: Theoretical and observed m/z for metabolites detected by mass spectrometry imaging.
